# The Association between Psoriasis and Risk of ST-Elevation Myocardial Infarction among African American Patients

**DOI:** 10.1101/2025.02.14.25322317

**Authors:** Ghena Krdi, Kennedy Sparling, Mohammad Reza Movahed, Mehrtash Hashemzadeh, Mehrnoosh Hashemzadeh

## Abstract

**Background/Objectives:** Several studies have attempted to investigate the association between psoriasis and the incidence of cardiovascular disease (CVD) including ST-segment elevation myocardial infarction (STEMI), possibly making psoriasis a risk factor for the development of CVD. African American (AA) patients have a higher prevalence of cardiovascular disease risk factors, representing a racial health disparity. There are very limited studies on the association between psoriasis and STEMIs among this specific patient population. This study aims to explore if an association exists between a diagnosis of psoriasis and the incidence of STEMI among AA patients.

**Methods:** Patients with specific ICD-10 codes for psoriasis and STEMI were included from the National Inpatient Sample (NIS) data. The rates of STEMI in patients with an established diagnosis of psoriasis were calculated and compared between AA and non-AA patients. An additional multivariate analysis was performed to adjust for CVD risk factors including age, gender, hypertension, hyperlipidemia, smoking, and diabetes.

**Results:** The analysis revealed no significant association between a diagnosis of psoriasis and STEMI incidence among AA patients. However, there was a significant positive association between psoriasis and STEMIs with p-value <0.001 among the total population despite race. After adjusting for confounding variables, there remained no significant association between psoriasis and STEMI among AA patients, and there was a significant negative association between psoriasis and STEMI among non-AA patients.

**Conclusion:** These findings suggest that psoriasis is not directly associated with STEMI among AA and non-AA patients, especially when controlling for confounding risks. Given the low prevalence of psoriasis among AA patients and the large burden of CVD in this patient population, further research will need to be conducted possibly guiding early CVD risk assessment and psoriasis management in AA patients.

## Introduction

Psoriasis is a chronic autoimmune skin condition classically marked by the development of scaly plaques, affecting approximately 2.2% to 3.15% of the United States population^1^. The ongoing interplay of various pro-inflammatory pathways contributes to the proliferation of keratinocytes and the disturbance of normal cell differentiation, resulting in the characteristic psoriasis skin lesions^2^. Unfortunately, the effects of this inflammatory response have been suggested to extend beyond the skin to affect various other organ systems. Psoriasis has been linked to a variety of comorbidities, including obesity, diabetes, dyslipidemia, and metabolic syndrome^3^. Given that these conditions are widely recognized risk factors for cardiovascular disease (CVD), it is understandable how many studies have highlighted an association between psoriasis and CVD incidence^4^. Notably, a 2022 meta-analysis evaluating the relationship between psoriasis and myocardial infarctions (MIs) found a relative risk of 1.23 for the European population and 2.17 for the East Asian population^5^. Similarly, a 2023 prospective cohort study displayed an association between CVD and psoriasis, with an odds ratio of 2.12. However, when accounting for conventional risk factors, the association between these two conditions decreased, with the odds ratio now at 1.19^6^. Considering the significant morbidity and mortality associated with these diseases, further evaluation of the relationship between psoriasis and CVD is warranted.

The variation in risk across demographics in the previously mentioned 2022 meta-analysis underscores the importance of further investigation into the relationship between psoriasis and MIs, particularly among potentially vulnerable populations. Notably, the African American (AA) population confronts a disproportionate burden when it comes to CVD and its associated risk factors. In fact, the 2021 REasons for Geographic And Racial Differences in Stroke (REGARDS) study highlighted this disparity, noting a hazard ratio of 1.42 for CVD events when comparing AA and white participants after adjusting for sociodemographic factors^7^. Moreover, an additional study comparing clinical outcomes in patients who received fibrinolysis for a ST-elevation myocardial infarction (STEMI) found a significant difference in 5-year mortality rates between AA and white patients. Specifically, AA patients exhibited a higher mortality rate, with an adjusted hazard ratio of 1.63^8^. These findings emphasize the critical need for further exploration into CVD events in AA patients as well as factors that may be associated, such as psoriasis.

Additionally, while there is extended literature on the relationship between CVD and psoriasis in white adults, there is a paucity of studies regarding this association in AA patients. A possible explanation for this includes the finding that AAs have the lowest prevalence of psoriasis of any other race/ethnicity. In fact, white individuals have been shown to have almost twice the odds of having psoriasis when compared to AA patients^9^. With the elevated odds of CVD in the AA population, this trend reversal seen with psoriasis further highlights the importance of evaluating the general relationship between CVD events and psoriasis. Furthermore, the lack of information regarding this relationship in the AA population underscores the need for further investigation into this topic.

Therefore, this study aims to explore the association between psoriasis diagnosis and STEMI prevalence among AA patients as compared to the general population. In doing so, the overarching goal is to further understand this complex relationship in AA patients in order to work toward fostering equitable health outcomes and addressing healthcare disparities contributing to the morbidity and mortality of these devastating conditions.

## Methods

We used the National Inpatient Sample (NIS) database for collecting our data and formulating a sample population for the study. The NIS is considered the largest publicly available database of inpatient care data from many hospitals across the United States that can be used for conducting clinical studies requiring large sample sizes. It was developed for the Healthcare Utilization Project (HCUP) and sponsored by the Agency for Healthcare Research and Quality (AHRQ).

We utilized the International Classification of Diseases, Tenth Revision (ICD-10) codes for psoriasis between the years 2016-2020 summarized in Table 1. After compiling the codes, the NIS data was used to gather a sample size of patients with these codes indicating an established diagnosis of psoriasis. Inclusion criteria was the following: patients with a psoriasis diagnosis based on the ICD-10 codes listed in Table 1. Exclusion criteria was the following: patients without an established psoriasis diagnosis. Race (including White, Black, Hispanic, Asian/Pacific Islander, Native American, and others) was also derived from the data and used to identify patients who are of African American descent, to be included in the data analysis. Data on the prevalence of the following conditions was also obtained: STEMI, hypertension, hyperlipidemia, smoking, and diabetes. Other data regarding age and gender were also obtained.

**Table 1.**
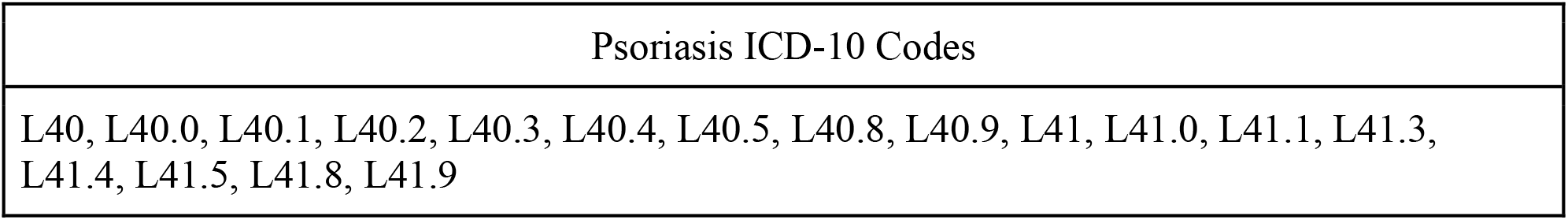
List of psoriasis ICD-10 codes for the years 2016-2020.

Using the Stat software, we performed a univariate analysis with the chi-square test to evaluate the odds ratio between psoriasis and all other collected data. Patient groups were divided into two groups: a general population group which includes patients from all races including AAs, and an AA population group which only includes the results from AA patients. Then, we conducted a multivariate analysis adjusting for risk factors of CVD including hypertension, hyperlipidemia, smoking and diabetes. Odds ratios (OR) and 95% confidence intervals (95% CI) were calculated. Two-sided P-values less than or equal to 0.05 were considered as statistically significant.

## Results

This study included a total of 174,776,205 patients from the NIS database between 2016 - 2020. Of those, 829,230 had a diagnosis of psoriasis based on the ICD-10 codes and were included in the study. Mean age 60.88±15.88 years, gender distribution 49.70% male and 50.28% female.

Race: 79.32% White, 5.52% Black (45,750 patients), 7.14% Hispanic, 2.23% Asian/Pacific Islander, 0.61% Native American, and 5.18% Others. CVD risk factors: 1.38% STEMI, 66.08% hypertension, 43.17% hyperlipidemia, 31.27% smoking, and 33.30% diabetes.

From the data, 45,750 patients identified as AA with an established diagnosis of psoriasis, which is equivalent to 5.52% of patients with an established psoriasis diagnosis. Mean age 57.13±16.39 years, gender distribution 43.33% male and 56.57% female. CVD risk factors: 1.20% had a STEMI, 73.83% hypertension, 36.31% hyperlipidemia, 26.33% smoking, and 38.95% diabetes.

Based on this univariate analysis, there is a significant positive association between psoriasis and STEMI in the general population (OR 1.12, P-value <0.001) while no significant association was found in the AA patients population (OR 1.14, P-value 0.18). Regardless, psoriasis was significantly positively associated with all of the studied CVD risk factors in both groups as shown in Table 2 (P-values <0.001).

**Table 2.** Prevalence of comorbidities among the total population of AA patients as compared to AA patients with an established diagnosis of psoriasis, P-values, ORs, and CIs.

|  | Total AA Population | AA with Psoriasis | P-value | OR (95% CI) |
| --- | --- | --- | --- | --- |
| STEMI | 1.06% | 1.20% | 0.18 | 1.14(0.94-1.38) |
| Hypertension | 52.53% | 73.84% | <0.001 | 2.55(2.43-2.68) |
| Hyperlipidemia | 24.38% | 36.31% | <0.001 | 1.77(1.69-1.85) |
| Smoking | 17.52% | 26.33% | <0.001 | 1.68(1.60-1.77) |
| Diabetes | 28.09% | 38.95% | <0.001 | 1.63(1.56-1.71) |

Multivariate analysis of the same data adjusting for the studied CVD risk factors (hypertension, hyperlipidemia, smoking, diabetes), summarized in Table 3, shows that there is a significant negative association between psoriasis and STEMIs among the general population (OR 0.88, p-value <0.001) which differs from the results shown in the univariate analysis. Similarly, psoriasis and STEMI maintained no significant association among AA patients after conducting the multivariate analysis (OR 0.87, p-value 0.17).

**Table 3.** OR and CI interval representing correlation between STEMI and psoriasis among general population group as compared to AA patients group.

| Population group | p-value | OR | OR (95% CI) |  |
| --- | --- | --- | --- | --- |
|  |  |  | Lower | Upper |
| General population | <0.001 | 0.88 | 0.84 | 0.92 |
| African American patients | 0.17 | 0.87 | 0.72 | 1.06 |

## Discussion

Our study aimed to investigate the association between psoriasis and STEMIs among AA patients as compared to the general population. Our results show a clinically significant association of increased STEMI prevalence among psoriasis patients from the general population (OR 1.12, P-value <0.001), which is otherwise not clinically significant from the AA patients group (OR 1.14, P-value 0.18). After performing a multivariate analysis adjusting for multiple cardiovascular risk factors, the results show a clinically significant decreased likelihood of STEMIs among psoriasis patients from the general population group (OR 0.88, p-value <0.001) and non-significant decreased likelihood from the AA patients group (OR 0.87, p-value 0.17). These results suggest that a direct association between psoriasis and STEMIs is not necessarily present.

Multiple studies were published on psoriasis and its link with CVD (including coronary artery disease, MIs, and atherosclerosis) ranging from meta-analysis to randomized trials. While no causal relationship between the two disease groups has been established, the ACC/AHA includes psoriasis as a risk-enhancing factor on its prevention of cardiovascular disease list^10^. Among the studies, some have shown a positive correlation or an increased relative risk of CVDs among psoriasis patients, while other studies only found such a relationship under certain conditions, or at times limited by confounding factors. Some of the explanations for these studies suggest that an established psoriasis association with CVD represents a dose-dependent relationship influenced by the duration, severity, and age of onset of psoriasis^10,11^. While our study does not take into account these measures, this relationship of positive association between these two conditions is consistent with our results from the univariate analysis.

An important aspect of our study focuses on identifying this relationship within AA patients. Our results did not show a clinically significant relationship between psoriasis and the prevalence of STEMI in this patient population both in the univariate and after conducting the multivariate analysis. A possible explanation for these findings may be linked to the prevalence of psoriasis among AA patients. A 2021 JAMA study that assessed the prevalence of psoriasis in different ethnic groups found that black patients had the lowest prevalence at 1.5% as compared to white patients which were the most prevalent at 3.6%^9^. An earlier study reported the prevalence of psoriasis among AA patients to be 4.8% in the United Kingdom to an absence in the United States, based on data published in 2014^12^. The same study found that psoriasis without arthritis was twice as frequent and more severe among AA patients [X]. Our NIS data sample of 829,230 patients with an established psoriasis diagnosis, 5.52% were black as compared to 79.32% white. The low prevalence might have affected the possibility of finding a significant relationship in the univariate analysis. Based on our research, our study is the first to investigate if a relationship exists between psoriasis and STEMIs among this patient population. Given the burden of cardiovascular disease and the mortality rate among AA patients, it is important to further study this relationship and identify ways to address this health disparity.

## Limitations

This study results can be affected by the sensitivity and specificity of the ICD-10 codes used to select patients. Similarly, under-diagnosis or over-diagnosis of psoriasis can affect the results. Furthermore, the inpatient sample may not be fully representative of the general population.

Some measures such as age of onset, severity, duration, and joint involvement of disease were not incorporated into the study.

## Conclusion

Based on a large inpatient database, the relationship between psoriasis and STEMI prevalence among the general population was positively clinically significant as compared to AA patients which was positive yet non-significant. After adjusting for cardiovascular risk factors including smoking, hyperlipidemia, HTN, and DM, the multivariate analysis no longer showed a clinically significant relationship among the general population. While our study focuses on investigating this relationship, future studies can investigate the interaction of genetics, severity, duration, age at onset, and joint involvement and how each of these correlate with CVD and its risk factors.

Some studies have already attempted some of these measures, but the current data is still limited.

## Data Availability

NIS database is publicly available

## Disclosure of conflict of interest

Ghena Krdi, MD: no conflicts to declare

Kennedy Sparling, MD: no conflicts to declare

Mohammad Reza Movahed, MD: no conflicts to declare

Mehrtash Hashemzadeh, MS: no conflicts to declare

Mehrnoosh Hashemzadeh, PhD: no conflicts to declare

Our institution does not require ethics approval for reporting [type of research, e.g., case study, secondary analysis, etc].

